# Family functioning but not social capital is associated with better mental health in adolescents affected by violence and displacement by armed conflict in Colombia

**DOI:** 10.1101/2021.06.24.21259443

**Authors:** William Tamayo-Aguledo, Alida Acosta-Ortiz, Aseel Hamid, Carolina Gómez-García, María Camila García-Durán, Vanessa Daccach-González, Francesca Solmi, Vaughan Bell

## Abstract

**Background:** The effect of the Colombian armed conflict on the mental health of adolescents is still poorly understood. Given social interventions are most likely to inform policy, we tested whether two potential intervention targets, family functioning and social capital, were associated with mental health in Colombian adolescents, and whether this was moderated by experience of violence and displacement.

**Methods:** We examined the cross-sectional association between family functioning, cognitive social capital, structural social capital and 12-month prevalence of Composite International Diagnostic Interview (CIDI) diagnosed psychiatric disorder, using data on 12-17-year-old adolescents (N = 1754) from the 2015 National Mental Health Survey of Colombia, a nationally representative epidemiological study. We tested whether associations survived cumulative adjustment for demographic confounders, experience of non-specific violence and harm, and displacement by armed conflict.

**Results:** Neither structural nor cognitive social capital were associated with better mental health. Better family functioning was associated with reduced risk of poor mental health in an unadjusted analysis (OR 0.90 [0.85 – 0.96]), and after cumulative adjustments for demographic confounders (OR 0.91 [0.86 – 0.97]), non-specific violence and harm (OR 0.91 [0.86 – 0.97]) and social capital variables (OR 0.91 [0.85 – 0.97]). In the final model, each additional point on the family APGAR scale was associated with a 9% reduced odds of any CIDI diagnosed disorder in the last 12 months.

**Conclusions:** Better family functioning was associated with better mental health outcomes for all adolescents. This effect remained present in those affected by the armed conflict even after accounting for potential confounders.

## Introduction

The Colombian armed conflict has endured for over five decades and despite a recent peace process, conflict-related violence has affected, and still affects, large numbers of people. One of the most marked effects has been the widespread impact of violence and the internal displacement of people. Estimates for those displaced by violence vary from 5 million (Internal Displacement Monitoring Centre, 2020) to 8 million people (Unidad para la Atención y la Reparación Integral a las Víctimas, 2021). The mental health of victims of the armed conflict and displaced people in Colombia has been identified as a priority (Chaskel et al., 2015) and is now codified in law. Nevertheless, the impact of armed conflict on the mental health of adolescents has been particularly under-researched meaning our understanding of how displacement affects adolescent mental health is still poorly understood (Tamayo-Agudelo and Bell, 2019).

Using data from the 2015 National Mental Health Survey of Colombia (NMHS; a large epidemiological study aiming for a representative sample of the population through stratified sampling) Marroquín Rivera et al (2020) reported that 5.3% of Colombian adolescents were displaced by violence, had higher rates of mental disorder, suicidal ideation and suicide attempts, and a greater prevalence of living below the poverty line in displaced compared to non-displaced participants. These results echo findings from smaller scale studies on displaced Colombian adolescents. In a sample of 471 displaced adolescents and young adults in three major cities, Sánchez Acosta et al. (2019) reported a Composite International Diagnostic Interview (CIDI) diagnosed prevalence of any mental disorder in the last 12 months of 24.4%. In a cross-sectional study, Gaias et al. (2019) reported that both the experience and witnessing of community violence were associated with externalizing behaviours in adolescents.

Notably, several of these studies also reported that factors relating to social support were associated with better mental health outcomes in adolescents affected by violence and displacement cross-sectionally. Sánchez-Villegas et al. (2021) reported family functioning and social support was associated with fewer emotional and behavioural problems, Sánchez Acosta et al. (2019) reported the same factors reduced the odds of CIDI diagnosed mental health problems, suicidal behaviour and substance use, and Gaias et al. (2019) reported positive school climate reduced the risk of externalising behaviours. Using the 2015 NMHS dataset, Gómez-Restrepo et al. (2021) reported that family dysfunction and economic disadvantage was associated with increased self-recognition of mental health problems in Colombian adolescents.

These factors are not solely of academic interest. Social and community interventions have been highlighted as an essential component for improving mental health in communities affected by armed conflict (Cummings et al., 2017; Jordans et al., 2013; World Health Organization, 2013) including in Colombia (Aranguren Romero and Rubio Castro, 2018; Burgess and Fonseca, 2020; MinSalud-ITES, 2017). However, currently there is a paucity of evidence that might help direct interventions and suggest how to focus them more effectively.

Two important components of social support have been hypothesised to be key in moderating poor mental health in those affected by conflict: social capital (Noel et al., 2018) and family functioning (Scharpf et al., 2021). Social capital relates to “features of social life — networks, norms, and trust — that enable participants to act together more effectively to pursue shared objectives” (Putnam, 1996). Within social capital, a common distinction is made between structural social capital – that refers to the extent and quantity of participation in social networks – including membership of groups, involvement in communal activities and support from individuals, and cognitive social capital – that describes the perception of the quality of the relationships, including perceptions related to trust, mutual help, and reciprocity (Harpham, 2008). Family functioning refers to the capacity of the family to provide emotional support and acceptance, resolve conflicts, remain cohesive in the face of challenges and communicate effectively (Alderfer et al., 2008).

Although researchers have debated to what extent different aspects of social capital and family functioning overlap (Almedom, 2005), they are measured as separate constructs, and importantly, at the policy level, suggest different approaches to intervention. Social capital interventions might involve community level interventions that help improve communication and trust within communities, encourage social groups that implicitly or explicitly provide social support, defuse tensions and bridge community divides, enable collective decision-making, or even shape the physical environment to facilitate social integration (Villalonga-Olives et al., 2018). Family interventions, on the other hand, may involve community level support for families, but are also likely to have a strongly clinical therapeutic focus that involves identifying and engaging conflictual families or those struggling to provide support to family members specifically to improve the mental health of the children or family members as a whole (Charlés, 2015).

Given the paucity of evidence for the effects of social capital and family functioning on mental health in adolescents and the potential moderating effects of violence and conflict both internationally and within Colombia, here we report an analysis of epidemiological data from the 2015 National Mental Health Survey of Colombia to address these questions. Specifically, we examined the association between family functioning, cognitive social capital and structural social capital and poor mental health, and tested whether any resultant associations would survive adjustment for demographic confounders, displacement by violence and armed conflict, and experience of non-specific violence and harm. Importantly, due to the fact that those living in areas controlled by armed groups in Colombia may be prohibited from engaging with or compelled to join formal social groups (Gáfaro et al., 2014) we distinguish structural social capital in terms of membership of formal community groups, and structural social capital involving availability of support from friends, partner, neighbours and other informal acquaintances.

## Methods

### Sample

The 2015 National Mental Health Survey (NMHS; in Spanish *La Encuesta Nacional de Salud Mental*) was a cross-sectional epidemiological study completed by the Ministry and Health and Social Protection (*Ministerio de Salud y Protección Social*) in Colombia. Data was collected using stratified sampling, aiming to be nationally representative of the population of Colombia, and included adults, adolescents and children. This was achieved by stratifying the population by region (Central, Oriental, Atlántica, Pacífica y Bogotá), then by municipality, then by geographical area. With each area, a section was selected (neighbourhood block in urban areas, and municipality sub-area in rural areas), and all households were contacted for participation. This study used data from the adolescent sample (N=1754) who were defined as being between 12-17 years old. Full details of the protocol and sampling methodology are reported in Gómez-Restrepo et al. (2016). Informed consent was obtained from all participants in the original epidemiological study and the study was approved by the review board of Pontificia Universidad Javeriana. This study is a secondary data analysis using the following data extracted from the total dataset.

### Measures

#### Mental health outcome

Participants were assessed using the electronic version of the Spanish-language Composite International Diagnostic Interview (CIDI-CAPI 3.0, henceforth referred to as CIDI; Rodriguez et al., 2016). The outcome used in this study was any CIDI diagnosed adolescent disorder during the last 12 months. It was coded as present if the individual fulfilled the diagnostic criteria for any one of major depressive disorder, dysthymia, mania, hypomania, generalised anxiety disorder, panic disorder or social phobia.

#### Displacement by armed conflict, violence and insecurity

Displacement by armed conflict was recorded in two items in the National Mental Health Survey. The first asked if the participant indicated earlier in the interview that they had moved residence (“Did any of the aforementioned changes of residence happen because your life or that of someone in your household was threatened by violence?”). In addition, an additional item asked “What was the main reason you changed your residence on the last occasion?” with one option being ‘armed conflict or insecurity’. If the participant reported on either of these items that they moved residence due to violence, they were coded as being displaced by violence. These items had a high proportion of “don’t know” answers (68.02%) but given the high salience of displacement by armed conflict, following past studies using this dataset (Gómez-Restrepo et al., 2021; Marroquín Rivera et al., 2020), we coded these as ‘no’ answers indicating no strong evidence of displacement by armed conflict.

#### Experience of non-specific violence and harm

Participants indicated whether they had been present, been a victim, or someone close to you had recounted they had been a victim of i) drowning, ii) an explosion or fire, iii) street robbery or mugging. These items were summed to create an overall score for exposure to non-specific violence and harm.

#### Social capital

The NMHS survey includes items drawn from Short Social Capital Assessment Tool (Harpham, 2008) and adapted for the Colombian context (Minsalud Colciencas, 2016) allowing us to calculate scores for cognitive social capital, structural social capital – support; and structural social capital – group membership.

The cognitive social capital variable was created by summing six items that represent perception of trust in the community and participation in the community for mutual benefit.

The structural social capital – support variable was created by summing six items each of which was a non-exclusive option to answer the question “From the following people, select those with whom you could discuss your problems or provide support if you need it”. Options included members of your family, friends, neighbours, partner, colleague from work or education, other, and the variable was created by summing the number of options selected by participants.

The structural social capital – group membership variable was created by summing 10 items. These items represented answers to the question “In which of the following groups do you participate?” and could be answered non-exclusively and included religious groups, sport groups, political groups, cultural groups, community groups, ecological groups, profession-based groups, ethnic groups, youth groups and health groups.

The original Spanish-language items and the English language translations for all social capital variables are included in Tables S1-S3 of the supplementary material.

#### Family functioning

Family functioning was measured with the family APGAR scale (Smilkstein, 1978), using the version translated and validated in Colombia (Ariza et al., 2006). This is a 5-item scale that measures an individual’s satisfaction with family relationships. It asks about satisfaction in terms of being able to turn to family for help, the way the family discusses and shares problems, accepts and supports the individual, expresses affection and responds to emotions, and the way the family share time together. Responses are coded from 0-4 (never, almost never, sometimes, almost always, always) and the total score was used in this study.

#### Demographics

The following demographic characteristics were analysed for each participant: age, sex, region, rural or urban living, and minority ethic status. Individuals were coded as living in poverty using the multidimensional poverty index (Alkire et al., 2014; Angulo et al., 2016) with those coded as scoring an urban or rural poverty index of 4, coded as living in a state of poverty.

#### Current neighbourhood security

Current neighbourhood security was rated by participants on a scale of 0 to 100, with 0 indicating the least safe possible and 100 indicating the safest neighbourhood possible.

### Analysis

The analysis plan was pre-registered and the pre-registration document for secondary data analysis is available at the following link: https://osf.io/k8bc2/

We planned a sequence of logistic regression models to examine to what extent social capital and family functioning was associated with any CIDI diagnosed adolescent psychiatric disorder during the last 12 months, after being adjusted for a potential confounders in a series of steps. These models were:

> Model 1: Are structural social capital – support, structural social capital –group membership, cognitive social capital and family functioning associated with any CIDI diagnosed disorder in the last 12 months?
>
> Model 2: Are associations in Model 1 explained by demographic confounders? Model 3: Are associations in Model 2 explained by displacement by armed conflict?
>
> Model 4: Are associations in Model 3 explained by non-specific violence and harm?
>
> Model 5: Are associations in Model 4 explained by inclusion of the other social capital / family functioning variables (i.e. is the original variable showing a unique association)?

Models 1-4 aimed to test whether structural capital and family functioning variables would be associated with CIDI diagnosed psychiatric disorder in the last 12 months after adjustment for demographic confounders, armed conflict, and general violence exposure.

Model 5 aimed to test whether these effects, if present, might be specific to the type of social capital / family functioning being measured. If the association is still present in Model 4 but is not present in Model 5, this will suggest that the association is non-specific. We renamed the variable originally described as ‘individual structural social capital’ in the pre-registration as ‘structural social capital – support’ to avoid confusion and better distinguish it from the ‘structural social capital – group membership’, as although it focuses on support available to the individual, it includes informal groups such as family members and friends.

## Results

All analysis was conducted using *R* version 4.0.3 (R Core Team, 2020) and were conducted on a Linux x86_64 platform. The *R* analysis code and calculated results are available as a Jupyter Notebook (Rule et al., 2019), a document that combines both code and the output in a form that can be re-run and reproduced, at the following link: https://osf.io/k8bc2/ Descriptive statistics for the sample and key study variables are displayed in Tables 1 and 2.

**Table 1.** Descriptive statistics

| *Variable* | *N (%)* | *Variable* | *Mean (SD)* |
| --- | --- | --- | --- |
| Sex (male) | 847 (48.3%) | Age | 14.5 (1.71) |
| Ethnic minority | 299 (17.0%) | Current neighbourhood security | 65.4 (29.5) |
| Living urban | 1362 (77.7%) |  |  |
| Living in poverty | 696 (39.7%) |  |  |
| Experience of |  |  |  |
| Drowning | 31 (1.8%) |  |  |
| Explosion or fire | 20 (1.1%) |  |  |
| Street robbery, mugging | 200 (11.4%) |  |  |
| Displacement by violence | 104 (5.9%) |  |  |
| Region |  |  |  |
| Central | 330 (18.8%) |  |  |
| Atlántica | 412 (23.5%) |  |  |
| Bogotá | 300 (17.1%) |  |  |
| Oriental | 411 (23.4%) |  |  |
| Pacífica | 301 (17.2%) |  |  |

**Table 2.** Means and standard deviations of social capital and family functioning stratified by prevalence of any CIDI diagnosed adolescent disorder during the last 12 months

|  | *12 month CIDI diagnosed disorder* |  | *Overall* |
| --- | --- | --- | --- |
|  | *Yes* | *No* |  |
| Structural social capital – support | 1.21 (0.618) | 1.39 (0.841) | 1.22 (0.64) |
| Structural social capital – group membership | 0.563 (0.745) | 0.558 (0.680) | 0.56 (0.74) |
| Cognitive social capital | 3.60 (1.55) | 3.71 (1.62) | 3.61 (1.55) |
| Family functioning (family APGAR score) | 17.3 (3.65) | 17.6 (3.28) | 17.3 (3.63) |

In terms of missing data, all variables had full data except for the following. The original items asking about the reason for the last move of residence had high rates of “don’t know” answers (68%). Following previous studies using this dataset (Gómez-Restrepo et al., 2021; Marroquín Rivera et al., 2020) we extracted responses that specifically indicated displacement by violence, insecurity or conflict as indicating displacement, meaning the recoded variable had no missing data. The items recording cognitive social capital had low frequencies of missing data (all less than 0.68%). The family APGAR scale was not present for 43.5% of participants due to the fact that the scale was only completed when the participant indicated that they had family ties after being given the following definition: “family is defined as the individual(s) with whom you usually live. If you live alone, your family corresponds to the person(s) with whom you have the strongest emotional ties at this time”. After being given this definition, N = 763 (43.5%) participants indicated they did not have relevant family ties. We note that this apparently high level of denial of family ties is concordant with some other results in the survey where N = 693 (39.5%) adolescents reported that their father did not live with them and N = 283 (16.1%) reported that their mother did not live with them, potentially indicating a high level of adolescents living in non-nuclear family situations. However, to account for this, we subsequently completed unplanned analyses to examine the effect of the absence of family ties, and to examine whether the results were substantially different when only those who reported family ties were included.

We also ran an unplanned analysis to examine the overall effect of structural social capital by combining ‘structural social capital – support’ and ‘structural social capital – group membership’ into a single structural social capital variable.

The results from the planned regression analyses are displayed in Table 3. Notably, none of the social capital variables were associated with mental health outcomes in either the unadjusted or adjusted analyses.

**Table 3.** Results of preregistered logistic regression analyses testing the association between social capital and family functioning variables and any CIDI diagnosed adolescent disorder during the last 12 months. Each cell reports the relevant odds ratio and 95% confidence intervals in brackets.

|  | *Model 1* | *Model 2* | *Model 3* | *Model 4* | *Model 5* |
| --- | --- | --- | --- | --- | --- |
|  | Unadjusted | + Demographic confounders | + Displacement by armed conflict | + Non-specific violence & harm | + Remaining social capital / family factors |
| Structural social capital – support | 1.32 (0.99 – 1.75) | 1.25 (0.93 – 1.68) | 1.22 (0.90 – 1.64) | 1.20 (0.89 – 1.62) | 1.45 (0.98 – 2.13) |
| Structural social capital – group membership | 0.82 (0.60 – 1.14) | 0.83 (0.60 – 1.14) | 0.83 (0.60 – 1.15) | 0.83 (0.60 – 1.14) | 1.06 (0.72 – 1.58) |
| Cognitive social capital | 0.88 (0.77 – 1.01) | 0.99 (0.85 – 1.14) | 0.98 (0.84 – 1.14) | 0.98 (0.85 – 1.14) | 0.97 (0.79 – 1.19) |
| Family functioning | 0.90 (0.85 – 0.96) | 0.91 (0.86 – 0.97) | 0.91 (0.86 – 0.97) | 0.91 (0.86 – 0.97) | 0.91 (0.85 – 0.97) |

In contrast, greater positive family functioning was associated with a reduced odds of any CIDI diagnosed disorder in the last 12 months in the unadjusted analysis, and also in subsequent analyses adjusted for potential demographic confounders, displacement by armed conflict, non-specific violence and harm, and the other social capital variables. In the final model, for each additional point on the family APGAR scale there was a 9% reduced odds of any CIDI diagnosed disorder in the last 12 months.

This suggest greater positive family functioning is associated with better mental health outcomes for all adolescents. This association was present irrespective of demographics, displacement by armed conflict, current neighbourhood safety, or experience of non-specific violence and harm.

In addition, we ran two unplanned analysis to account for the high rate of participants who reported a lack of family ties, and therefore did not have a family APGAR score. In the first, we examined the effect of lack of family ties on outcomes and after adjustment for confounders. Lack of family ties was not significantly associated with mental health in an unadjusted analysis (OR = 0.71 [0.45 – 1.11], p = 0.131), and after sequential adjustment for demographic confounders (OR = 0.70 [0.44 – 1.11], p = 0.125), displacement by armed conflict (OR = 0.69 [0.43 – 1.10], p = 0.119) and non-specific violence and harm (OR = 0.69 [0.43 – 1.10], p = 0.118).

In the second, we re-ran the full set of models but only included adolescents who reported family ties. The pattern of results remained the same with the exception that ‘structural social capital – support’ became significantly associated with an increased odds of poor mental health in the unadjusted analysis (OR = 1.50 [1.06 - 2.12], p = 0.023), and after sequential adjustment for demographic confounders (OR = 1.54 [1.06 - 2.25], p = 0.025), displacement by armed conflict (OR = 1.48 [1.01 - 2.18], p = 0.043), and non-specific violence and harm (OR = 1.48 [1.01 - 2.18], p = 0.044). Cognitive social capital was associated with reduced odds of poor mental health in the unadjusted analysis (OR = 0.81 [0.69 - 0.97], p = 0.019) but was not associated with mental health in any of the adjusted analyses. Full results are reported in Table S3 in the supplementary material and in the online archive.

We also completed an additional unplanned analysis to examine the association between a total social structural capital score (created by summing structural social capital – support, and structural social capital – group membership variables) and outcome. Total structural capital was not significantly associated with mental health in an unadjusted analysis (OR = 1.03 [0.84 – 1.27], p = 0.746), and after sequential adjustment for demographic confounders (OR = 1.01 [0.82 – 1.24], p = 0.91), displacement by armed conflict (OR = 1.00 [0.81 – 1.23], p = 0.99) and non-specific violence and harm (OR = 0.99 [0.80 – 1.22], p = 0.93).

## Discussion

We examined cross-sectional associations between family functioning, cognitive and structural social capital, and poor mental health in adolescents in a representative epidemiological study of Colombia. We examined whether associations found in the initial analysis survived adjustment by demographic confounders, displacement by violence and armed conflict, and experience of non-specific violence and harm. Consequently, to examine whether any particular social factors made a unique contribution to mental health outcome, we subsequently adjusted for the remaining social factors. Our analysis showed that better family functioning was reliably associated with lower odds of poor mental health after being fully adjusted for demographics, displacement and violence, suggesting that this may be a protective factor. In contrast, social capital variables were not reliably associated with better mental health across multiple analyses.

The results indicate that for each additional point on the family APGAR scale there was a 9% reduced odds of psychiatric disorder in the last 12 months. This study was cross-sectional and so the causal role of family functioning in reducing adolescent mental health problem cannot be confidently inferred from the data reported here. Family functioning has been associated with better mental health in children and adolescents in cross-sectional studies (Scully et al., 2020) with longitudinal studies suggesting a bidirectional effect of family functioning and adolescent mental health (Breaux and Harvey, 2019; Shek, 1998) indicating that better family functioning may improve adolescent mental health but that poor adolescent mental health may also impact negatively on family functioning. This evidence suggests that it is plausible that at least some of the effect reported here does reflect the causal role of family functioning as a protective factor for adolescent mental health and might be a feasible target for intervention.

This raises the question as to what extent this effect is meaningful in terms of a potential guide to prioritising social interventions – in this case, family interventions for adolescents affected by displacement and violence. Indeed, if this was a fully causal contribution and family APGAR score accurately reflected family functioning, a 9% reduction in outcome per increased point would be a substantial contribution to improving adolescent mental health both at the family level and at the population level. We also note that that any potential benefits for mental health are likely to be in addition to other social, economic and health benefits of supporting positive family functioning for displaced adolescents (Denov et al., 2019; Scharpf et al., 2021).

However, it is worth sounding a note of caution. Although APGAR scores covered the full range of possible scores, the distribution was right-skewed with the majority of adolescents reporting high levels of positive family functioning. In contrast, a high level of adolescents (over 40%) did not complete the family APGAR because they indicated on the screening question that they did not have relevant family ties corresponding to the definition of a family as ‘the people with whom you usually live or, if living alone, the people with whom you have the strongest emotional ties’. Although subsequent analyses attempted to take account of this and showed substantial difference in those with and without family ties this may reflect measurement issues. We note here that the family APGAR has been validated in Colombia (Ariza et al., 2006), but that family functioning was reported self-reported by adolescents who may have been hesitant to admit problems in their family due to a belief that family issues should remain private (Keitner and Miller, 1990). Indeed, under-reporting may be a particular issue for those affected by the Colombian armed conflict where an attempt to conceal difficulties may be a mechanism for attempting to avoid further perceived harm to the family (Grupo de Memoria Histórica, 2013).

It is also important to highlight that associations between family functioning and mental health outcomes in displaced children and adolescents have varied across countries (Reed et al., 2012) and it is likely that outcomes are a result of an interaction between exposure to violence, parental factors, and cultural models of what ‘good family functioning’ entails (d’Abreu et al., 2019). Indeed, this is likely to vary across social contexts in Colombia and over time given the diversity of cultures within the country and the changing dynamic of the conflict (Bell et al., 2012; Cifuentes Patiño, 2009). Nevertheless, evidence from this study indicates an overall association with better mental health in adolescents, with evidence to suggest that some of this contribution may be causal. However, further work is needed to test this more thoroughly and to investigate whether there are specific factors that moderate these associations.

It is also worth noting the association between ‘structural social capital – support’, and increased odds of poor mental health. This association was not present in the planned analyses but was present when we re-ran the analysis to only include those with family ties to examine whether those with family ties differed in terms of potential benefits of social capital. In contrast to any benefits potential, we found it was associated with worse mental health in those with existing family ties. Structural social capital measures have shown a mixed relationship with mental health outcomes in low income countries including being associated with an increased risk for poor mental health (De Silva et al., 2007). Indeed, similar mixed results have been reported in Colombia, where social capital has been found to be associated with better mental health (Cardozo Rusinque et al., 2017), not associated with it (Harpham et al., 2004) or associations have been reported that vary in direction depending on the nature of social capital being measured and identified groups (Hurtado et al., 2011). Notably, a study in post-conflict Guatemala (Dinesen et al., 2013) found that structural social capital was a risk factor for victimisation by violence, potentially suggesting that even if it is a protective factor that reduces the risk of mental health problems post-violence, it may also increase the risk of being exposed to violence.

We also note some important limitations to our study. First, the analysis was completed on cross-sectional data meaning it is hard to confidently determine the direction of causality. Notably, we restricted the outcome to ‘any CIDI diagnosed adolescent psychiatric disorder during the last 12 months’ to attempt to reduce the chances of reverse causality by making it more likely that the outcome occurred after other measured events. This was most likely for displacement by violence, insecurity and armed conflict, assuming, in line with the profile of the displaced population of Colombia in that the majority of displaced people sampled were unlikely to have been displaced within the last 12 months of the long-running conflict. It is possible that better family functioning was an effect of displacement, rather than a moderator of outcome, although given what we know about the impact of displacement on families, we think this is unlikely in most cases (Shultz et al., 2014).

Parental mental health problems have been identified as a risk factor for poor outcome for adolescent mental health (Van Loon et al., 2014), which was something that was not addressed in this study. Although parental mental health problems have been hypothesised to impact adolescent mental health through their impact on family functioning (Leinonen et al., 2003), it remains possible that the unmeasured effect of parental mental health led to residual confounding. It is also worth noting that the social capital items were drawn from an established measure (Harpham, 2008) and were adapted to the Colombian context for the national mental health survey. Nevertheless, although they closely match the items from the original scale, they have not been validated in their adapted form.

We also note that the rate of displacement by violence reported by adolescents was 5.9% although the original items asking about reasons for the last change of residence had high rates of “don’t know” responses. However, this estimate is broadly in line with prior estimates of the demographics of displaced people in Colombia, where 11.6% of the population have been internally displaced and approximately half are under 18 years of age (Consuelo Carrillo, 2009).

In conclusion, we report that family functioning was associated with reduced odds of poor mental health in Colombian adolescents and this remained true when adjusting for demographic confounders, displacement by violence and armed conflict, and experience of non-specific violence and harm. However, we note some of the questions we raise about potential directions of causality are poorly addressed by cross-sectional data. This highlights the need for cohort studies to better understand potential causal influences on mental health as these are currently lacking in people affected by the Colombian armed conflict, and indeed, in people affected by armed conflict globally.

## Data Availability

The data is available on application to the Ministerio de Salud y Proteccion Social of Colombia

## Acknowledgements

Thank you to Professor Carlos Gómez-Restrepo and Consul Josè Ricardo Puyana for assistance with the application process for obtaining the data used in this study.

## References

Alderfer, M.A., Fiese, B.H., Gold, J.I., Cutuli, J.J., Holmbeck, G.N., Goldbeck, L., Chambers, C.T., Abad, M., Spetter, D., Patterson, J., 2008. Evidence-based Assessment in Pediatric Psychology: Family Measures. Journal of Pediatric Psychology 33, 1046–1061. https://doi.org/10.1093/jpepsy/jsm083

Alkire, S., Chatterjee, M., Conconi, A., Seth, S., Vaz, A., 2014. Global multidimensional poverty index 2014, OPHI Briefing 21. University of Oxford, Oxford.

Almedom, A.M., 2005. Social capital and mental health: An interdisciplinary review of primary evidence. Social Science & Medicine 61, 943–964. https://doi.org/10.1016/j.socscimed.2004.12.025

Angulo, R., Díaz, Y., Pardo, R., 2016. The Colombian Multidimensional Poverty Index: Measuring Poverty in a Public Policy Context. Soc Indic Res 127, 1–38. https://doi.org/10.1007/s11205-015-0964-z

Aranguren Romero, J.P., Rubio Castro, N., 2018. Formación en herramientas terapéuticas a sobrevivientes del conflicto armado en el Pacífico colombiano: reflexividad y cuidado de sí. Revista de Estudios Sociales 18–29.

Ariza, L.M.F., Durán, M.C.A., Cubillos, Z.J.D., Arias, A.C., 2006. Consistencia interna y análisis de factores de la escala APGAR para evaluar el funcionamiento familiar en estudiantes de básica secundaria. Revista colombiana de psiquiatría 35, 23–29.

Bell, V., Méndez, F., Martínez, C., Palma, P.P., Bosch, M., 2012. Characteristics of the Colombian armed conflict and the mental health of civilians living in active conflict zones. Conflict and Health 6, 10. https://doi.org/10.1186/1752-1505-6-10

Breaux, R.P., Harvey, E.A., 2019. A Longitudinal Study of the Relation Between Family Functioning and Preschool ADHD Symptoms. Journal of Clinical Child & Adolescent Psychology 48, 749–764. https://doi.org/10.1080/15374416.2018.1437737

Burgess, R.A., Fonseca, L., 2020. Re-thinking recovery in post-conflict settings: Supporting the mental well-being of communities in Colombia. Global Public Health 15, 200– 219. https://doi.org/10.1080/17441692.2019.1663547

Cardozo Rusinque, A.A., Cortés-Peña, O.F., Castro Monsalvo, M., 2017. Relaciones funcionales entre salud mental y capital social en víctimas del conflicto armado y personas en situación de pobreza. [Functional relationships between mental health and social capital in victims of armed conflict and people in poverty.]. Interdisciplinaria Revista de Psicología y Ciencias Afines 34, 235–257.

Charlés, L.L., 2015. Scaling Up Family Therapy in Fragile, Conflict-Affected States. Family Process 54, 545–558. https://doi.org/10.1111/famp.12107

Chaskel, R., Gaviria, S.L., Espinel, Z., Taborda, E., Vanegas, R., Shultz, J.M., 2015. Mental health in Colombia. BJPsych International 12, 95–97. https://doi.org/10.1192/S2056474000000660

Cifuentes Patiño, M.R., 2009. Familia y conflicto armado. Trabajo Social 87–106.

Consuelo Carrillo, A., 2009. Internal displacement in Colombia: humanitarian, economic and social consequences in urban settings and current challenges. Int’l Rev. Red Cross 91, 527.

Cummings, E.M., Merrilees, C.E., Taylor, L.K., Mondi, C.F., 2017. Developmental and social–ecological perspectives on children, political violence, and armed conflict. Development and Psychopathology 29, 1–10. https://doi.org/10.1017/S0954579416001061

d’Abreu, A., Castro-Olivo, S., Ura, S.K., 2019. Understanding the role of acculturative stress on refugee youth mental health: A systematic review and ecological approach to assessment and intervention. School Psychology International 40, 107–127. https://doi.org/10.1177/0143034318822688

De Silva, M.J., Huttly, S.R., Harpham, T., Kenward, M.G., 2007. Social capital and mental health: A comparative analysis of four low income countries. Social Science & Medicine 64, 5–20. https://doi.org/10.1016/j.socscimed.2006.08.044

Denov, M., Fennig, M., Rabiau, M.A., Shevell, M.C., 2019. Intergenerational resilience in families affected by war, displacement, and migration: “It runs in the family.” Journal of Family Social Work 22, 17–45. https://doi.org/10.1080/10522158.2019.1546810

Dinesen, C., Ronsbo, H., Juárez, C., González, M., Estrada Méndez, M.Á., Modvig, J., 2013. Violence and social capital in post-conflict Guatemala. Rev Panam Salud Publica 34, 162–168.

Gáfaro, M., Justino, P., Ibáñez, A.M., 2014. Collective action and armed group presence in Colombia. Documento CEDE.

Gaias, L.M., Johnson, S.L., White, R.M.B., Pettigrew, J., Dumka, L., 2019. Positive School Climate as a Moderator of Violence Exposure for Colombian Adolescents. American Journal of Community Psychology 63, 17–31. https://doi.org/10.1002/ajcp.12300

Gómez-Restrepo, C., de Santacruz, C., Rodriguez, M.N., Rodriguez, V., Tamayo Martínez, N., Matallana, D., Gonzalez, L.M., 2016. Encuesta Nacional de Salud Mental Colombia 2015. Protocolo del estudio. Revista Colombiana de Psiquiatría, Encuesta Nacional de Salud Mental 2015 45, 2–8. https://doi.org/10.1016/j.rcp.2016.04.007

Gómez-Restrepo, C., Malagón, N.R., Eslava-Schmalbach, J., Ruiz, R., Gil, J.F., 2021. Factores asociados al reconocimiento de trastornos y problemas mentales en adolescentes en la Encuesta Nacional de Salud Mental, Colombia. Revista Colombiana de Psiquiatría 50, 3–10. https://doi.org/10.1016/j.rcp.2019.09.002

Grupo de Memoria Histórica, 2013. ¡Basta ya! Colombia: memorias de guerra y dignidad: informe general. Centro Nacional de Memoria Histórica.

Harpham, T., 2008. The measurement of community social capital through surveys, in: Social Capital and Health. Springer, pp. 51–62.

Harpham, T., Grant, E., Rodriguez, C., 2004. Mental health and social capital in Cali, Colombia. Social Science & Medicine 58, 2267–2277. https://doi.org/10.1016/j.socscimed.2003.08.013

Hurtado, D., Kawachi, I., Sudarsky, J., 2011. Social capital and self-rated health in Colombia: The good, the bad and the ugly. Social Science & Medicine 72, 584–590. https://doi.org/10.1016/j.socscimed.2010.11.023

Internal Displacement Monitoring Centre, 2020. Global Report on Internal Displacement 2020. IDMC, Geneva.

Jordans, M.J.D., Tol, W.A., Susanty, D., Ntamatumba, P., Luitel, N.P., Komproe, I.H., Jong J.T.V.M. de, 2013. Implementation of a Mental Health Care Package for Children in Areas of Armed Conflict: A Case Study from Burundi, Indonesia, Nepal, Sri Lanka, and Sudan. PLOS Medicine 10, e1001371. https://doi.org/10.1371/journal.pmed.1001371

Keitner, G.I., Miller, I.W., 1990. Family functioning and major depression: An overview. The American Journal of Psychiatry 147, 1128–1137. https://doi.org/10.1176/ajp.147.9.1128

Leinonen, J.A., Solantaus, T.S., Punamäki, R.-L., 2003. Parental mental health and children’s adjustment: the quality of marital interaction and parenting as mediating factors. Journal of Child Psychology and Psychiatry 44, 227–241. https://doi.org/10.1111/1469-7610.t01-1-00116

Marroquín Rivera, A., Rincón Rodríguez, C.J., Padilla-Muñoz, A., Gómez-Restrepo, C., 2020. Mental health in adolescents displaced by the armed conflict: findings from the Colombian national mental health survey. Child and Adolescent Psychiatry and Mental Health 14, 23. https://doi.org/10.1186/s13034-020-00327-5

Minsalud Colciencas, 2016. Encuesta Nacional de Salud Mental 2015 Tomo 1. Minsalud Colciencas, Bogota. MinSalud-ITES, 2017. Protocolo de Atención Integral en Salud con Enfoque Psicosocial a Víctimas del Conflicto Armado. MinSalud, Bogota.

Noel, P., Cork, C., White, R.G., 2018. Social Capital and Mental Health in Post-Disaster/Conflict Contexts: A Systematic Review. Disaster Medicine and Public Health Preparedness 12, 791–802. https://doi.org/10.1017/dmp.2017.147

Putnam, R.D., 1996. The strange disappearance of civic America. Policy: A Journal of Public Policy and Ideas 12, 3–15.

R Core Team, 2020. R: A language and environment for statistical computing. R Foundation for Statistical Computing, Vienna, Austria.

Reed, R.V., Fazel, M., Jones, L., Panter-Brick, C., Stein, A., 2012. Mental health of displaced and refugee children resettled in low-income and middle-income countries: risk and protective factors. The Lancet 379, 250–265. https://doi.org/10.1016/S0140-6736(11)60050-0

Rodriguez, V., Moreno, S., Camacho, J., Gómez-Restrepo, C., de Santacruz, C., Rodriguez, M.N., Tamayo Martínez, N., 2016. Diseño e implementación de los instrumentos de recolección de la Encuesta Nacional de Salud Mental Colombia 2015. Revista Colombiana de Psiquiatría, Encuesta Nacional de Salud Mental 2015 45, 9–18. https://doi.org/10.1016/j.rcp.2016.10.001

Rule, A., Birmingham, A., Zuniga, C., Altintas, I., Huang, S.-C., Knight, R., Moshiri, N., Nguyen, M.H., Rosenthal, S.B., Pérez, F., Rose, P.W., 2019. Ten simple rules for writing and sharing computational analyses in Jupyter Notebooks. PLOS Computational Biology 15, e1007007. https://doi.org/10.1371/journal.pcbi.1007007

Sánchez Acosta, D., Castaño Pérez, G.A., Sierra Hincapié, G.M., Semenova Moratto Vásquez, N., Salas Zapata, C., Buitrago Salazar, J.C., Torres de Galvis, Y., Sánchez Acosta, D., Castaño Pérez, G.A., Sierra Hincapié, G.M., Semenova Moratto Vásquez, N., Salas Zapata, C., Buitrago Salazar, J.C., Torres de Galvis, Y., 2019. Salud mental de adolescentes y jóvenes víctimas de desplazamiento forzado en Colombia. CES Psicología 12, 1–18. https://doi.org/10.21615/cesp.12.3.1

Sánchez-Villegas, M., Reyes-Ruiz, L., Taylor, L.K., Pérez-Ruíz, N.A., Carmona-Alvarado, F.A., 2021. Mental health problems, family functioning and social support among children survivors of Colombia’s armed conflict. Journal of Aggression, Conflict and Peace Research 13, 61–72. https://doi.org/10.1108/JACPR-08-2020-0535

Scharpf, F., Kaltenbach, E., Nickerson, A., Hecker, T., 2021. A systematic review of socio-ecological factors contributing to risk and protection of the mental health of refugee children and adolescents. Clinical Psychology Review 83, 101930. https://doi.org/10.1016/j.cpr.2020.101930

Scully, C., McLaughlin, J., Fitzgerald, A., 2020. The relationship between adverse childhood experiences, family functioning, and mental health problems among children and adolescents: a systematic review. Journal of Family Therapy 42, 291–316. https://doi.org/10.1111/1467-6427.12263

Shek, D.T.L., 1998. A Longitudinal Study of the Relationship between Family Functioning and Adolescent Psychological Well-Being. Journal of Youth Studies 1, 195–209. https://doi.org/10.1080/13676261.1998.10593006

Shultz, J.M., Garfin, D.R., Espinel, Z., Araya, R., Oquendo, M.A., Wainberg, M.L., Chaskel, R., Gaviria, S.L., Ordóñez, A.E., Espinola, M., Wilson, F.E., García, N.M., Ceballos, Á.M.G., Garcia-Barcena, Y., Verdeli, H., Neria, Y., 2014. Internally Displaced “Victims of Armed Conflict” in Colombia: The Trajectory and Trauma Signature of Forced Migration. Curr Psychiatry Rep 16, 475. https://doi.org/10.1007/s11920-014-0475-7

Smilkstein, G., 1978. The family APGAR: a proposal for a family function test and its use by physicians. J fam pract 6, 1231–9.

Tamayo-Agudelo, W., Bell, V., 2019. Armed conflict and mental health in Colombia. BJPsych International 16, 40–42. https://doi.org/10.1192/bji.2018.4

Unidad para la Atención y la Reparación Integral a las Víctimas, 2021. Registro Único de Víctimas (RUV) [WWW Document]. Unidad para las Víctimas. URL https://www.unidadvictimas.gov.co/es/registro-unico-de-victimas-ruv/37394 (accessed 6.8.21).

Van Loon, L.M.A., Van de Ven, M.O.M., Van Doesum, K.T.M., Witteman, C.L.M., Hosman, C.M.H., 2014. The Relation Between Parental Mental Illness and Adolescent Mental Health: The Role of Family Factors. J Child Fam Stud 23, 1201–1214. https://doi.org/10.1007/s10826-013-9781-7

Villalonga-Olives, E., Wind, T.R., Kawachi, I., 2018. Social capital interventions in public health: A systematic review. Social Science & Medicine 212, 203–218. https://doi.org/10.1016/j.socscimed.2018.07.022

World Health Organization, 2013. World Health Organization mental health action plan 2013–2020. WHO, Geneva.

