## Supplementary material for "Family functioning but not social capital is associated with better mental health in adolescents affected by violence and displacement by armed conflict in Colombia"

| *NMHS Item* | *Original question in Spanish* | *English translation* |
| --- | --- | --- |
| m2_p57 | ¿La mayoría de sus vecinos están dispuestos a ayudar cuando otro vecino tiene una emergencia? | Are most of your neighbours willing to help out when another neighbour has an emergency? |
| m2_p58 | ¿Puede confiar en la mayoría de sus vecinos? | Can you trust most of your neighbours? |
| m2_p59 | Si un proyecto de la comunidad no lo beneficia directamente, pero tiene beneficios para muchas otros vecinos, ¿usted contribuiría? | If a community project does not directly benefit you, but benefits many other neighbours, would you contribute? |
| m2_p60 | Si hubiera un problema con el suministro de agua o luz ¿usted y sus vecinos se ayudarían entre si? | If there was a problem with the water or electricity supply, would you and your neighbours help each other? |
| m2_p61 | ¿Usted o alguien de su hogar tiene como práctica frecuente participar en alguna actividad en beneficio de la comunidad? | Do you or someone in your household frequently participate in any activity for the benefit of the community? |
| m2_p61 | ¿Si a usted se le perdiera la billetera fuera de su casa, cree que se la devolverían? | If you lost your wallet outside your home, do you think it would be returned to you? |

**Table S1.** Summed items used to calculate the cognitive social capital variable

Prompt question:

De las siguientes personas, seleccione aquellas que con las que usted podría discutir sus problemas o le brindarían apoyo si lo necesitara

From the following people, select those with whom you could discuss your problems or provide support if you need it

| *NMHS Item* | *Original item in Spanish* | *English translation* |
| --- | --- | --- |
| m2_p55__1 | Miembros de su familia | Members of your family |
| m2_p55__2 | Amigos | Friends |
| m2_p55__3 | Vecinos | Neighbours |
| m2_p55__4 | Pareja | Partner |
| m2_p55__5 | Compañero de trabajo o de estudio | Colleague from work or education |
| m2_p55__6 | Otro | Other |

**Table S2.** Summed items used to calculate the individual structural social capital variable

Prompt question:

¿En cuáles de los siguientes grupos participa?

In which of the following groups do you participate?

| *NMHS Item* | *Original item in Spanish* | *English translation* |
| --- | --- | --- |
| m2_p56__1 | Grupos religiosos | Religious groups |
| m2_p56__2 | Grupos deportivos | Sport groups |
| m2_p56__3 | Grupos políticos | Political groups |
| m2_p56__4 | Grupos culturales | Cultural groups |
| m2_p56__5 | Grupos comunitarios | Community groups |
| m2_p56__6 | Grupos ecológicos | Ecological groups |
| m2_p56__7 | Grupos gremiales | Profession-based groups |
| m2_p56__8 | Grupos étnicos | Ethnic groups |
| m2_p56__9 | Grupos juveniles | Youth groups |
| m2_p56__10 | Grupos de salud | Health groups |

**Table S3.** Summed items used to calculate the group structural social capital variable

|  | *Model 1* |  | *Model 2* |  | *Model 3* |  | *Model 4* |  | *Model 5* |
| --- | --- | --- | --- | --- | --- | --- | --- | --- | --- |
|  | Unadjusted |  | + Demographic confounders |  | + Displacement by armed conflict |  | + Non-specific violence & harm |  | + Remaining social capital / family factors |
| Structural social capital – support | 1.50 (1.06 - 2.12) |  | 1.54 (1.06 - 2.25) |  | 1.48 (1.01 - 2.18) |  | 1.48 (1.01 - 2.18) |  | 1.45 (0.98 - 2.13) |
| Structural social capital – group membership | 1.02 (0.69 - 1.50) |  | 1.02 (0.70 - 1.51) |  | 1.02 (0.69 - 1.51) |  | 1.02 (0.69 - 1.51) |  | 1.06 (0.72 - 1.58) |
| Cognitive social capital | 0.81 (0.69 - 0.97) |  | 0.96 (0.79 - 1.17) |  | 0.95 (0.78 - 1.15) |  | 0.98 (0.85 - 1.14) |  | 0.97 (0.79 - 1.19) |
| Family functioning | 0.90 (0.85 - 0.96) |  | 0.91 (0.86 - 0.97) |  | 0.91 (0.86 - 0.97) |  | 0.91 (0.86 - 0.97) |  | 0.91 (0.85 - 0.97) |

**Table S4.** Results of all analyses repeated to include only those adolescents who reported family ties. Each cell reports the relevant odds ratio and 95% confidence intervals in brackets. The analysis for family functioning and Model 5 are identical as they depend on the presence of a family APGAR score as per the prior analyses but are included for ease of comparison.
